# Altered CSF Tryptophan–Kynurenine Pathway Metabolism in Multiple System Atrophy: Distinct from Classic Microglial Markers

**DOI:** 10.1101/2025.07.16.25331609

**Authors:** Ryunosuke Nagao, Kazuya Kawabata, Yasuaki Mizutani, Sayuri Shima, Akihiro Ueda, Mizuki Ito, Yasuhiro Maeda, Akihiro Mouri, Hirohisa Watanabe

**Affiliations:** Department of Neurology, Fujita Health University School of Medicine, Toyoake, Aichi, Japan; Division of BrainTheraInformatics, International Center for Brain Science (ICBS), Fujita Health University, Toyoake, Aichi, Japan; Fujita Mind-Brain Research & Innovation Center for Drug Generation (Fujita Mind-BRIDGe); Department of Neurology, Fujita Health University Okazaki Medical Center, Okazaki, Aichi, Japan; Department of Neurology, Fujita Health University Bantane Hospital, Nagoya, Aichi, Japan; Open Facility Center, Fujita Health University, Toyoake, Aichi, Japan; Department of Regulatory Science for Evaluation & Development of Pharmaceuticals & Devices, Fujita Health University Graduate School of Medical Sciences, Toyoake, Aichi, Japan; Division of Neurochemistry, ICBS, Fujita Health University, Toyoake, Aichi, Japan

**Keywords:** Multiple system atrophy, Kynurenine, Kynurenic acid, Quinolinic acid, Tryptophan

## Abstract

**Background:** Alterations in tryptophan (TRP)-serotonin metabolism have been implicated in multiple system atrophy (MSA). However, the involvement of the TRP-kynurenine (KYN) pathway, which is associated with neuroinflammatory and neuroprotective processes, is elusive.

**Objectives:** To investigate tryptophan metabolism in cerebrospinal fluid (CSF), we examined the relationships between metabolites and their associations with clinical biomarkers reflecting microglial activation and axonal injury in MSA patients.

**Methods:** CSF and clinical data from 51 patients with clinically established MSA and 56 control subjects were analyzed. CSF concentrations of TRP, KYN, 3-hydroxykynurenine (3-HK), quinolinic acid (QA), and kynurenic acid (KA) were quantified. Additionally, 5-hydroxyindoleacetic acid (5-HIAA), glycoprotein nonmetastatic melanoma protein B (GPNMB), soluble triggering receptor expressed on myeloid cells 2 (sTREM2), and neurofilament light chain (NfL) were measured. Group differences and inter-biomarker correlations were assessed, with hierarchical cluster analysis performed on the resulting correlation matrix. Principal component analysis (PCA) was performed to explore the latent relationships among biomarkers.

**Results:** Patients with MSA showed higher CSF levels of QA, a neuroinflammatory marker (adjusted median difference controlling for age and sex, 2.56 nM; 95% CI, 0.99–3.60) and lower levels of KA, which is considered neuroprotection (−6.46 nM; 95% CI, −9.08 to −3.12), yielding an elevated QA/KA ratio (+0.52; 95% CI, 0.27–0.88). In contrast, CSF levels of TRP, KYN, and 3-HK did not differ significantly between the two groups. TRP was correlated only with KYN (Spearman’s ρ = 0.56, adjusted for age and sex), while KYN was correlated with 3-HK (ρ = 0.47) and QA (ρ = 0.64), forming a shared cluster. KA did not show significant correlations with any of the measured kynurenine metabolites. Although GPNMB, sTREM2, and NfL were also increased, these inflammatory and neurodegenerative markers showed no correlation with kynurenic metabolites and formed distinct clusters. PCA indicated that these markers are independent of KYN pathway metabolites.

**Conclusions:** This study revealed a marked imbalance in KYN metabolism in MSA, which suggests a metabolic shift that may promote excitotoxicity and proinflammatory processes that are distinct from classic neuroinflammatory markers. These findings may highlight future therapeutic strategies targeting the kynurenine pathway.

## Introduction

Multiple system atrophy (MSA) is a sporadic, adult-onset neurodegenerative disorder characterized by varying degrees of autonomic dysfunction, cerebellar ataxia, and parkinsonism (Wenning *et al*., 2022). The progression of symptoms varies among patients, with rapid deterioration of clinical features accompanied by pronounced atrophic changes (Watanabe *et al*., 2002; Krismer *et al*., 2023; Kawabata *et al*., 2025). A major pathological hallmark of MSA is the presence of α-synuclein-positive glial cytoplasmic inclusions that are accompanied by cell loss and demyelination (Wakabayashi *et al*., 1998). Furthermore, pathological neuroinflammation has attracted increasing interest as a key feature of MSA and may contribute to disease progression (Vieira *et al*., 2015; Hoffmann *et al*., 2019; Leńska-Mieciek *et al*., 2023).

Tryptophan (TRP) is metabolized via two major pathways: the serotonin (5-HT) pathway and the kynurenine (KYN) pathway, with the latter implicated in neuroinflammation and neuroprotection. We previously reported that the TRP-5-HT pathway metabolite 5-hydroxyindoleacetic acid (5-HIAA) was significantly reduced in the cerebrospinal fluid (CSF) of patients with MSA and was correlated with disease severity (Nagao *et al*., 2024). In TRP metabolism, only a small fraction of TRP is converted into 5-HT, whereas approximately 95% is catabolized by the KYN pathway (Bender, 1983; Haq *et al*., 2021; Badawy and Guillemin, 2022). 5-HT synthesis is suppressed during TRP depletion, which is mediated by proinflammatory cytokines that induce indoleamine 2,3-dioxygenase 1 (IDO1), thereby increasing KYN pathway activity (Haq *et al*., 2021). KYN is metabolized by kynurenine 3-monooxygenase (KMO) into 3-hydroxykynurenine (3-HK), which generates endogenous oxidative stress, and further downstream, quinolinic acid (QA), which is an endogenous agonist of the N-methyl-D-aspartate (NMDA) receptor (Okuda *et al*., 1998). KYN is also metabolized by kynurenine aminotransferases (KATs) into kynurenic acid (KA), an endogenous antagonist of the NMDA receptor (Moroni *et al*., 2012; Schwarcz *et al*., 2012). QA is thus recognized for its neurotoxic and proinflammatory properties, whereas KA is considered neuroprotective. Accordingly, the QA/KA ratio is considered a surrogate marker of neuroinflammatory status (Mor *et al*., 2021; Kaleta *et al*., 2024). While TRP-KYN pathway dysregulation in neuroinflammation has been reported in neurodegenerative disorders such as Alzheimer’s disease (AD) and Parkinson’s disease (PD) (Sorgdrager *et al*., 2019; Choe *et al*., 2025; Wilson *et al*., 2025), its role in MSA–a distinct α-synucleinopathy with a more aggressive disease course–has not been characterized. Given MSA’s unique glial pathology and rapid progression, investigating the KYN pathway may reveal disease-specific metabolic signature that are not evident in other neurodegenerative conditions.

Therefore, based on our previous findings of alterations in the 5-HT pathway and the established role of neuroinflammation in MSA, we hypothesized that TRP-KYN pathway metabolites are altered in the brain and CSF. We further investigated their associations with other CSF biomarkers of microglial activation and neurodegeneration in MSA, including glycoprotein nonmetastatic melanoma protein B (GPNMB), soluble triggering receptor expressed on myeloid cells 2 (sTREM2), and neurofilament light chain (NfL).

## Methods

### Participants

This study included 51 patients with clinically established MSA, comprising 25 MSA-cerebellar subtypes (MSA-C) and 26 MSA-parkinsonian subtypes (MSA-P), who were diagnosed according to the diagnostic criteria of the International Parkinson and Movement Disorder Society (MDS) for MSA (Wenning *et al*., 2022) and 56 control subjects (Table 1). All participants were free from serotonergic medications, which may affect tryptophan metabolism. MSA symptoms and severity were assessed using the Unified MSA Rating Scale (UMSARS). Cognitive performance was assessed using the Japanese version of the Montreal Cognitive Assessment (MoCA-J), the Frontal Assessment Battery (FAB), the Mini-Mental State Examination (MMSE), and the Addenbrooke’s Cognitive Examination-Revised (ACE-R). Nonmotor symptoms were assessed using the Geriatric Depression Scale-15 (GDS-15) for depression, the Odor Stick Identification Test for Japanese (OSIT-J) for olfactory function, and the Scales for Outcomes in Parkinson’s Disease–Autonomic (SCOPA-AUT) for autonomic symptoms. This study was approved by the Ethics Committee of Fujita Health University Hospital (HM23-296). Written informed consent, including the provision for opt-out, was obtained from all participants prior to their inclusion. This study conformed to the principles described in the Declaration of Helsinki.

**Table 1.** Demographics and clinical characteristics of patients with MSA and controls.

| Variable | MSA<br>n = 51 | Control<br>n = 56 | P-value |
| --- | --- | --- | --- |
| Age, y | 63.3 ± 9.5 | 63.6 ± 11.3 | 0.884 |
| Sex, female/male, n | 27/24 | 20/36 | 0.082 |
| MSA-P/MSA-C, n | 26/25 | - | NA |
| Disease duration, m | 30.3 ± 24.3 | - | NA |
| UMSARS Part I | 16.5 ± 9.9 | - | NA |
| UMSARS Part II | 19.8 ± 9.3 | - | NA |
| UMSARS Part III |  | - | NA |
| SBP drop, mmHg | 29.3 ± 19.3 | - | NA |
| DBP drop, mmHg | 12.5 ± 11.8 | - | NA |
| HR change, bpm | 6.5 ± 6.2 | - | NA |
| UMSARS Part IV | 2.3 ± 1.2 | - | NA |
| MMSE | 27.1 ± 3.3 | - | NA |
| MoCA-J | 22.0 ± 4.7 | - | NA |
| ACE-R | 86.7 ± 12.9 | - | NA |
| FAB | 14.1 ± 2.6 | - | NA |
| GDS-15 | 8.0 ± 3.9 | - | NA |
| OSIT-J | 8.9 ± 2.6 | - | NA |
| SCOPA-AUT | 15.5 ± 6.8 | - | NA |

Data are presented as mean ± standard deviation (SD) for continuous variables and as counts for categorical variables (sex). P-values were calculated using the chi-square test for sex and Welch's t-test for age.

### CSF Collection and Analytes

For the MSA group, CSF samples were prospectively collected via a standard lumbar puncture performed after fasting for at least 8 hours. Within 2 hours of collection, the samples were centrifuged at 1500 × g for 10 minutes at 4 °C, and 500 µL aliquots of the supernatant were transferred into polypropylene tubes and stored at −80 °C until analysis. For the present study, stored CSF samples were retrospectively retrieved and used for biomarker measurements. In contrast, owing to the invasive nature of lumbar puncture, CSF samples from neurologically healthy individuals were obtained during spinal anesthesia for benign urological procedures, with written informed consent, as part of an aging registry study approved by the institutional review board at Fujita Health University. CSF levels of TRP–KYN pathway metabolites, including TRP, KYN, 3-HK, QA, and KA, were measured. To investigate the associations between KYN metabolites and markers of microglial activity and axonal damage, the levels of GPNMB (a potential marker for neurodegeneration), sTREM2 (microglial activation), and NfL (axonal damage) were also measured. CSF protein and albumin levels were measured to estimate bloodCbrain barrier (BBB) dysfunction in the MSA group (Gabdulkhaev *et al*., 2025), and the albumin quotient (Qalb = [CSF albumin/serum albumin] × 1000) was calculated in same-day serum samples. To reference serotonergic involvement, available data on 5-HIAA for 49 MSAs and 14 controls were included.

### Measurement of CSF TRP, KYN and 3-HK

An internal standard mixture in MeOH was prepared as a mixture of 0.5 μM TRP d5 and 0.5 μM KYN d4. A CSF sample (15 μL) and the internal standard mixture (15 μL) were mixed, vortexed, and centrifuged (15,000 × *g for* 10 min at 4 °C). Three microliters of the supernatant was injected into a UHPLCCMS/MS system (LCMS-8060; Shimadzu, Kyoto, Japan). Analytes were eluted from a Discovery HS F5 column (150 × 2.1 mm, 3 μm; Supelco, Merck, St. Louis, MO) with the gradient method using a mobile phase with 0.1% aqueous formic acid solution and 0.1% formic acid in acetonitrile at a flow rate of 0.25 mL/min and were detected by multiple reaction monitoring mode of MS/MS.

### Measurement of CSF KA and QA

An internal standard mixture in EtOH was prepared as a mixture of 0.1 μM QA d3 and 1 μM KA d5. To 15 μL of cerebrospinal fluid, 15 μL of an internal standard mixture, 30 μL of 7.5% pyridine in 75% methanol, 30 μL of 50 mM 3-nitrophenylnydradine in 75% methanol, and 30 μL of 50 mM 1-ethyl-3-(3-dimethylaminopropyl) carbodiimide in 75% methanol were added. After the mixture was shaken at room temperature for 30 min, 60 μL of 5% formic acid in 75% methanol was added. The mixture was subsequently centrifuged (15,000 × *g* for 10 min at 4 °C), after which the supernatant was injected into a UHPLC-MS/MS instrument (LCMS-8060NX; Shimadzu, Kyoto, Japan). Analytes were eluted from a reversed-phase column (InertSustain AQ-C18, 150 × 2.1 mm, 1.9 μm; GL Sciences, Tokyo, Japan) with the gradient method using a mobile phase with 0.1% aqueous formic acid solution and acetonitrile at a flow rate of 0.35 mL/min and were detected by the multiple reaction monitoring mode of MS/MS.

### Measurement of GPNMB, sTREM2, NfL

GPNMB, sTREM2, and NfL in CSF were measured using commercial ELISA kits (ab224881, Abcam, Cambridge, UK; DY2550, R&D Systems, Minneapolis, MN, USA; and NF-light™ ELISA RUO, UmanDiagnostics AB, Umeå, Sweden, respectively).

### Measurement of CSF 5-HIAA

CSF 5-HIAA levels were measured via high-performance liquid chromatography according to previously described methods (Mailman and Kilts, 1985; Nagao *et al*., 2024). CSF was stored at 4 °C after collection, and analysis was conducted within one day. The CSF 5-HIAA level was considered unchanged during the storage period on the basis of a previous study (Javors *et al*., 1984). The molecular weight of 5-HIAA was calculated to be 191 g/mol, and the measurement unit was standardized.

### Statistical analysis

Continuous variables such as age and clinical scores are presented as means ± standard deviations. Group differences in age were assessed using Welch’s t-test, and differences in sex distribution were evaluated using the chi-square test. In contrast, as several metabolite measures showed skewed distributions with outliers, nonparametric statistics were used for biomarkers, and the data are presented as medians and interquartile ranges (IQRs) on the basis of raw values. Therefore, group comparisons were performed using the Wilcoxon rank-sum test on unadjusted values, and median differences adjusted for age and sex were estimated using quantile regression without imputation for missing values. Ninety-five percent confidence intervals (95% CI) were calculated using bootstrap resampling with 10000 permutations. Receiver operating characteristic (ROC) curve analysis was conducted to evaluate the discriminative ability of QA/KA to distinguish between MSA patients and controls.

To explore the relationships among biomarkers and clinical scores in MSA patients, Spearman’s correlation analysis and principal component analysis (PCA) were conducted within the MSA group. In these analyses, because complete cases, also known as listwise deletions, yield biased estimates of associations (Pedersen *et al*., 2017), missing values were handled using multiple imputation with predictive mean matching by the *mice* package in R (version 4.5.0). A total of 100 imputed datasets were generated using an imputation model that incorporated all measured biomarkers, clinical scores, age, and sex under the assumption of missing at random (MAR), allowing for unbiased and valid estimation of associations on the basis of the observed data (Pedersen *et al*., 2017). Convergence of the imputation model was assessed using the potential scale reduction factor (PSRF), with further details provided in the supplementary materials. Subsequently, Spearman’s correlation analyses were conducted on each dataset, and the statistical measures were pooled using Rubin’s rules (Rubin, 1987; Marshall *et al*., 2009). To perform hierarchical clustering, correlation coefficients were converted into distance metrics using (1-Spearman correlation) with complete linkage, generating heatmaps using the *pheatmap* package in R. PCA including CSF biomarkers was conducted for each imputed dataset, and Procrustes alignment was applied to integrate the component structures across imputations, averaging both the component scores and loadings (van Ginkel, 2023). As a sensitivity analysis, we also performed correlation analysis using pairwise deletion and PCA using listwise deletion for missing values to assess the robustness of our findings without an imputation process. Given the exploratory nature of the study, unadjusted p values are reported. Additionally, to account for multiple comparisons, false discovery rate (FDR) correction was applied across simultaneously tested outcome variables, with q values < 0.05 considered statistically significant.

All the statistical analyses were conducted using JMP Pro 16.0 (SAS Institute Inc., Cary, NC, USA), GraphPad Prism, and R version 4.5.0.

## Results

The demographic and clinical characteristics of the 51 patients with MSA and the 56 controls are summarized in Table 1. There were no significant differences in sex or age between the MSA and control groups.

### Alterations in CSF Tryptophan Metabolites in MSA

Figure 1 and Table 2 illustrate alterations in the KYN pathway in MSA. Compared with controls, patients with MSA had significantly greater QA (median 7.77 in MSA vs. 4.94 nM in controls; adjusted median difference, +2.56 nM; 95% CI, 0.99–3.60) and lower KA (8.40 vs. 17.4 nM; −6.46 nM; 95% CI, −9.08 to −3.12), resulting in an elevated QA/KA ratio (0.81 vs. 0.27; +0.52; 95% CI, 0.27–0.88). CSF 5-HIAA was also reduced (11.4 in MSA patients vs. 18.1 ng/mL in controls; −6.58 ng/mL; 95% CI, −11.9 to −1.85). These metabolites remained significant after correction for multiple comparisons (FDR q < 0.05). No significant group differences were observed for TRP, KYN, 3-HK, or the KYN/TRP ratio. ROC analysis revealed that the QA/KA ratio distinguished MSA patients from controls, with an area under the curve (AUC) of 0.86 (95% CI, 0.79– 0.94); a cutoff of 0.52 yielded a sensitivity of 0.71 (95% CI, 0.55–0.92) and a specificity of 0.91 (95% CI, 0.70–1.00) (Figure 1B). No significant differences in CSF TRP metabolites were observed between the MSA-P and MSA-C subtypes (Supplementary Table S1).

**Figure 1.**
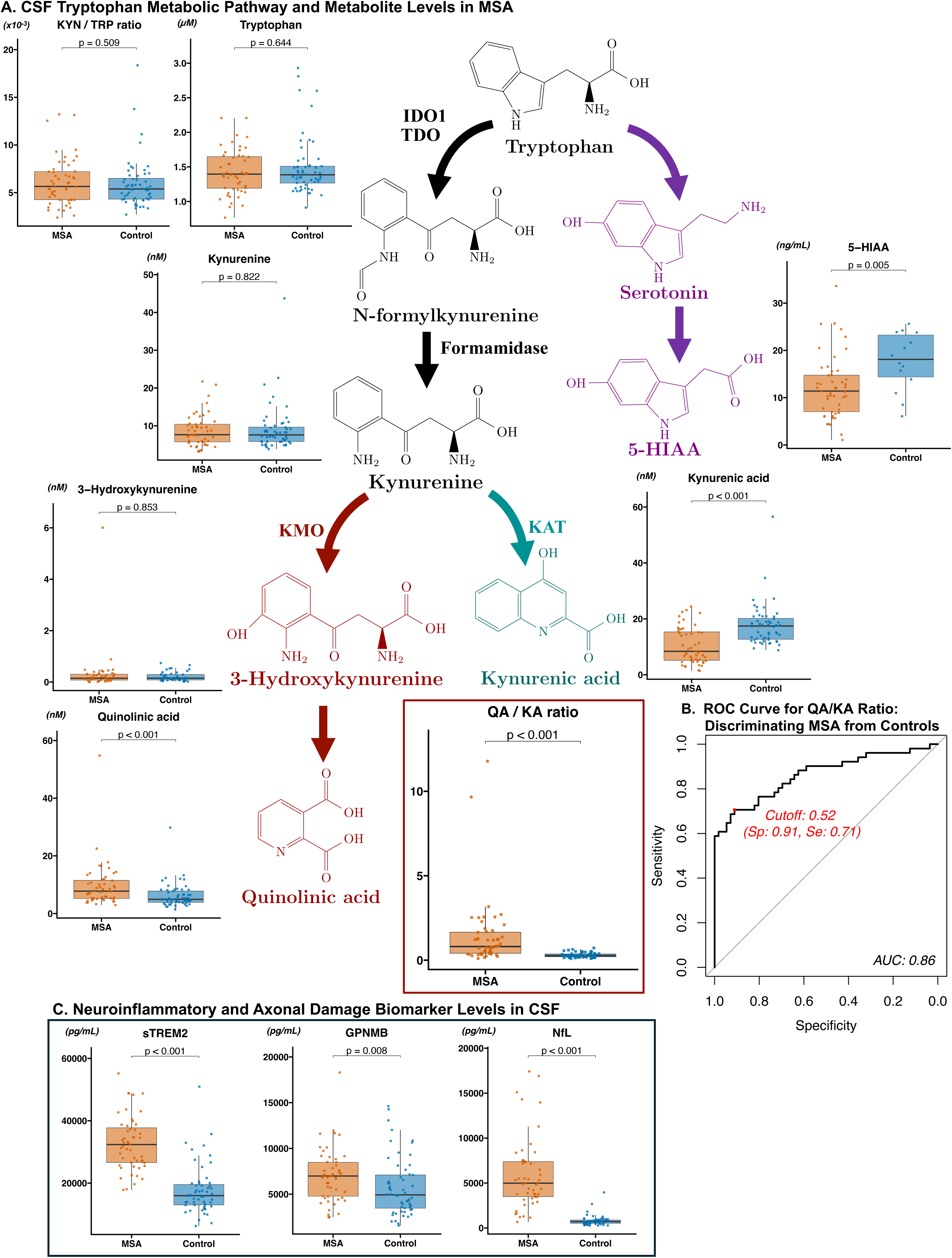
Overview of the CSF TRP Pathway and Related Biomarker Analyses in MSA. A. CSF tryptophan (TRP) metabolic pathway and its alterations in MSA. Major pathway metabolites and corresponding boxCandCdot plots are shown for TRP, kynurenine (KYN), the KYN/TRP ratio, 3-hydroxykynurenine, quinolinic acid (QA), kynurenic acid (KA), the QA/KA ratio, and 5-HIAA. Box plots represent the median and interquartile range. P values were calculated by the Wilcoxon rank-sum test. All p values less than 0.01 survived after controlling false discovery rate (FDR) q < 0.05. B. Receiver operating characteristic (ROC) curve for differentiating MSA patients from controls by the QA/KA ratio. C. Box-and-dot plots for CSF sTREM2, GPNMB, and NfL. 5-HIAA, 5-hydroxyindoleacetic acid; CSF, cerebrospinal fluid: GPNMB, glycoprotein nonmetastatic melanoma protein B; IDO1, indoleamine-2,3-dioxygenase 1; KAT, kynurenine amino transferase; KMO, kynurenine 3-monooxygenase; NfL, neurofilament light chain; sTREM2, soluble triggering receptor expressed on myeloid cells 2; TDO, tryptophan-2,3-dioxygenase.

**Table 2.**
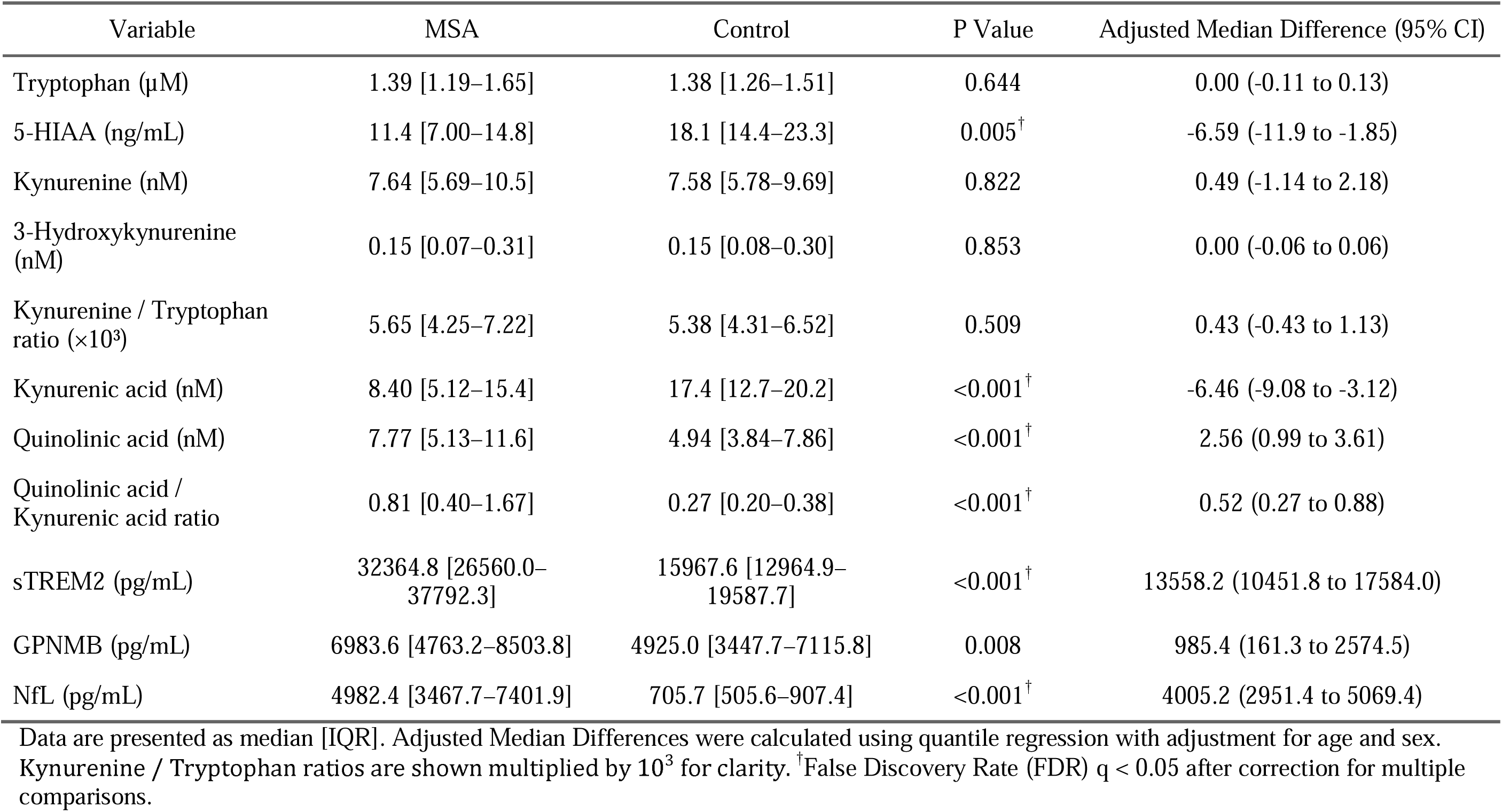
Tryptophan pathway metabolites and biomarker differences between MSA and Control.

| Variable | MSA | Control | P Value | Adjusted Median Difference (95% CI) |
| --- | --- | --- | --- | --- |
| Tryptophan (μM) | 1.39 [1.19–1.65] | 1.38 [1.26–1.51] | 0.644 | 0.00 (-0.11 to 0.13) |
| 5-HIAA (ng/mL) | 11.4 [7.00–14.8] | 18.1 [14.4–23.3] | 0.005<sup>†</sup> | -6.59 (-11.9 to -1.85) |
| Kynurenine (nM) | 7.64 [5.69–10.5] | 7.58 [5.78–9.69] | 0.822 | 0.49 (-1.14 to 2.18) |
| 3-Hydroxykynurenine (nM) | 0.15 [0.07–0.31] | 0.15 [0.08–0.30] | 0.853 | 0.00 (-0.06 to 0.06) |
| Kynurenine / Tryptophan ratio (×10<sup>3</sup>) | 5.65 [4.25–7.22] | 5.38 [4.31–6.52] | 0.509 | 0.43 (-0.43 to 1.13) |
| Kynurenic acid (nM) | 8.40 [5.12–15.4] | 17.4 [12.7–20.2] | <0.001<sup>†</sup> | -6.46 (-9.08 to -3.12) |
| Quinolinic acid (nM) | 7.77 [5.13–11.6] | 4.94 [3.84–7.86] | <0.001<sup>†</sup> | 2.56 (0.99 to 3.61) |
| Quinolinic acid / Kynurenic acid ratio | 0.81 [0.40–1.67] | 0.27 [0.20–0.38] | <0.001<sup>†</sup> | 0.52 (0.27 to 0.88) |
| sTREM2 (pg/mL) | 32364.8 [26560.0–37792.3] | 15967.6 [12964.9–19587.7] | <0.001<sup>†</sup> | 13558.2 (10451.8 to 17584.0) |
| GPNMB (pg/mL) | 6983.6 [4763.2–8503.8] | 4925.0 [3447.7–7115.8] | 0.008 | 985.4 (161.3 to 2574.5) |
| NfL (pg/mL) | 4982.4 [3467.7–7401.9] | 705.7 [505.6–907.4] | <0.001<sup>†</sup> | 4005.2 (2951.4 to 5069.4) |

Data are presented as median [IQR]. Adjusted Median Differences were calculated using quantile regression with adjustment for age and sex. Kynurenine / Tryptophan ratios are shown multiplied by 10<sup>3</sup> for clarity. <sup>†</sup>False Discovery Rate (FDR) q < 0.05 after correction for multiple comparisons.

### Neuroinflammatory and Neurodegenerative Biomarkers in CSF

The MSA group had higher CSF levels of sTREM2 (median 32365 in MSA vs. 15968 pg/mL in controls; adjusted median difference, +13558 pg/mL; 95% CI, 10452–17584), GPNMB (6984 vs. 4925 pg/mL; +985 pg/mL; 95% CI, 161–2575), and NfL (4982 vs. 706 pg/mL; +4005 pg/mL; 95% CI, 2951–5069), with these differences remaining significant after FDR adjustment (q < 0.05). In the control group, one sample each for sTREM2 and NfL was undetectable. No significant differences in these biomarkers were observed between the MSA-P and MSA-C subtypes (Supplementary Table S1).

### Relationships among CSF metabolites in the MSA group

Correlation analysis of CSF analytes adjusted for age and sex, along with hierarchical clustering based on the correlation matrix, is shown in Figure 2A. Hierarchical clustering revealed three distinct groups by visual inspection of the dendrogram: (1) TRP, KYN, 3-HK, and QA; (2) sTREM2, GPNMB, and NfL; and (3) KA and 5-HIAA. Within the KYN pathway, TRP was correlated with KYN (Spearman’s ρ = 0.56, p < 0.001), and KYN was correlated with 3-HK (ρ = 0.47, p = 0.002) and QA (ρ = 0.64, p < 0.001), all of which survived FDR correction (q < 0.05). KA did not show significant correlations with any of the measured kynurenine metabolites. In contrast, among the biomarkers of sTREM2 and GPNMB, correlations were observed between sTREM2 and GPNMB (ρ = 0.53, p < 0.001) and between GPNMB and NfL (ρ = 0.55, p < 0.001). None of the sTREM2, GPNMB, or NfL were associated with KYN, 3-HK, QA, or KA. Sensitivity analyses using unadjusted data and datasets without multiple imputation confirmed the robustness of the observed relationships among metabolites and biomarkers (Supplementary Figure S1).

**Figure 2.**
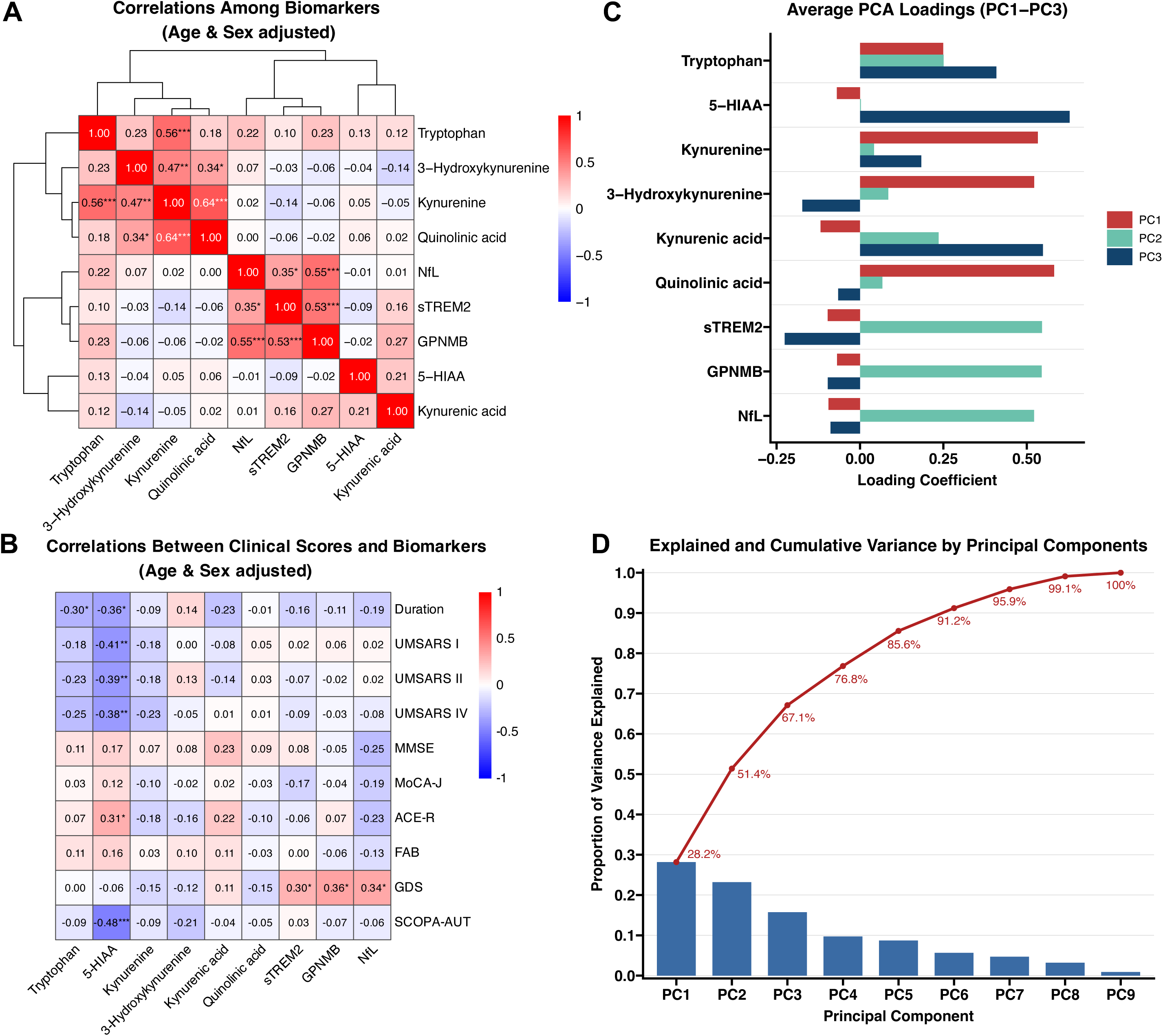
Relationships among Biomarkers, Clinical Features, and Principal Component Structures. A. Correlations among biomarkers adjusted for age and sex using quantile regression residuals. The values represent Spearman’s ρ. * p < 0.05; ** p < 0.01; *** p < 0.001. All p-values are provided in supplementary table S2. B. Correlations between clinical scores and biomarkers adjusted for age and sex. All p-values are provided in supplementary table S3. C. Average loadings from principal component analysis (PCA) for principal components (PCs) 1 to 3 across biomarkers. D. Explained variance and cumulative variance of the PCs. 5-HIAA, 5-hydroxyindoleacetic acid; GPNMB, glycoprotein nonmetastatic melanoma protein B; NfL, neurofilament light chain; sTREM2, soluble triggering receptor expressed on myeloid cells 2.

With respect to the relationships between clinical scores and CSF TRP-KYN pathway metabolites, no significant correlations were observed, except for a weak inverse correlation between disease duration and tryptophan. However, after applying FDR correction, none of the associations remained statistically significant. An association between 5-HIAA and autonomic and disease severity scores was observed, consistent with our previous findings (Nagao et al., 2024).

Given that QA and KA do not readily cross the BBB under normal conditions, we examined their associations with Qalb to evaluate the potential influence of BBB dysfunction. QA was not correlated with Qalb (ρ = 0.09, p = 0.532), whereas KA was negatively correlated with Qalb (ρ = –0.35, p = 0.011), indicating possible susceptibility to BBB-related loss (Supplementary Figure S3). Consequently, the QA/KA ratio was correlated with Qalb (ρ = 0.40, p = 0.003).

### Principal component analysis

The PCA results are shown in Figure 2C and 2D. Although the first five principal components explained more than 80% of the total variance, only the first three (each contributing >10% and collectively explaining 67%) were retained for downstream analysis to ensure interpretability, as the contributions of the fourth and fifth components were less than 10% (Figure 2D). PC1 exhibited high loadings for KYN, 3-HK, and QA, reflecting KYN pathway metabolism toward QA (Figure 2C). PC2 showed high loadings for sTREM2, GPNMB, and NfL, indicating the presence of neuroinflammatory and axonal injury markers. PC3 captured variability related to 5-HIAA and KA. TRP shows relatively balanced loading across components. PC loadings from PC1 to PC3 without multiple imputation are also shown in Supplementary Figure S1 and are largely consistent with the main analysis.

## Discussion

This study provides novel insights into alterations in CSF TRP-KYN pathway metabolism in patients with MSA. We identified a marked imbalance in KYN metabolites, with elevated levels of the neurotoxic metabolite QA and reduced levels of the neuroprotective KA, resulting in a high distinction in the ratio of QA/KA between MSA patients and controls. Our results also suggest a neuroinflammatory shift in TRP metabolism in MSA, independent of changes in other biomarkers, such as sTREM2, GPNMB, and NfL.

Dysregulation of the TRP-KYN pathway has been reported across a wide range of neuropsychiatric disorders in the brain. Both KA and QA have limited permeability across the BBB, making their local production within the brain particularly relevant. Astrocytes predominantly produce the neuroprotective metabolite KA, whereas microglia are the primary source of neurotoxic metabolites such as QA (Platten *et al*., 2019). Notably, the direction of metabolic imbalance in the brain differs between neurological and psychiatric conditions. While psychiatric disorders are associated with elevated KA levels (Mori *et al*., 2021; Hasegawa *et al*., 2025; Wang *et al*., 2025), neurodegenerative disorders typically show a shift toward reduced KA and increased QA (Wang *et al*., 2025). QA is an NMDA receptor agonist and neurotoxin implicated in excitotoxicity, oxidative stress, and neuroinflammation and is involved in several neurological disorders, such as AD, PD, Huntington’s disease, and ALS (Lugo-Huitrón *et al*., 2013; Huang *et al*., 2023). In contrast, KA exerts neuroprotective effects by blocking NMDA receptors and modulates cognition and mood through antagonism of the α7-nicotinic acetylcholine receptor (α7nAChR) (Platten *et al*., 2019).

In the KYN pathway, KMO is a key regulatory enzyme that modulates the balance between QA and KA production (Parrott and O’Connor, 2015). Located on the outer mitochondrial membrane, KMO catalyzes the conversion of KYN to 3-HK, thereby promoting the production of QA while reducing KA synthesis. Its activation is also associated with the generation of reactive oxygen species (ROS), mitochondrial dysfunction, and neuronal injury itself (Castellano-Gonzalez *et al*., 2019; Mor *et al*., 2021). In our study, QA levels were elevated in the CSF of patients with MSA, and the QA/KA ratio was markedly different between MSA patients and controls. The observed elevation in QA, a major neurotoxic endpoint of the KYN pathway with limited conversion to NAD^+^ in the brain, may reflect an increased downstream metabolic flux mediated by KMO, despite unchanged 3-HK levels, possibly due to its role as a transient intermediate metabolized by competing enzymes (Schwarcz *et al*., 2012). Although KMO activity was not directly assessed, the observed metabolite profile is consistent with chronic KMO upregulation, which may promote QA accumulation, forming a pathogenic cycle that exacerbates neuronal vulnerability (Hughes *et al*., 2022). Given KMO’s mitochondrial localization and its dual role in metabolism and cytotoxicity, targeting this KMO-associated metabolic imbalance may inform future research on therapeutic strategies in MSA. Our study focused on downstream metabolic signatures rather than upstream immunological drivers, which will be important to evaluate in future multiomics investigations. Future studies incorporating enzymatic assays or gene expression profiling of KMO and related enzymes are warranted.

Furthermore, the KYN-to-TRP ratio, a widely used surrogate marker of IDO1 activity, did not differ between MSA patients and controls. IDO1 is an inducible enzyme upregulated by proinflammatory cytokines, particularly interferon-γ (IFN-γ), under inflammatory conditions. In MSA, neuroinflammation is characterized by the activation of microglia and astrocytes, with IFN-γ being implicated as a potential upstream regulator of IDO1 expression (Schwarcz *et al*., 2012; Vieira *et al*., 2015). Given the preserved KYN/TRP ratio despite the presence of inflammation, significant induction of IDO1 appears unlikely. Instead, the KYN pool may be primarily maintained by tryptophan 2,3-dioxygenase (TDO), a constitutively expressed hepatic enzyme that regulates systemic KYN production under steady-state conditions. Although KYN can cross the BBB, its stable levels of KYN in CSF, along with a marked increase in QA, suggest that the metabolic shift toward neurotoxicity may be driven by increased KMO activity rather than IDO1 induction. Moreover, Qalb, an indicator of BBB integrity (Gabdulkhaev *et al*., 2025), was negatively correlated with KA but not with QA alone. This pattern, which is consistent with findings in an AD study (van der Velpen *et al*., 2019), indicates that while KA efflux may be increased in association with increased barrier permeability, the increase in QA itself is driven primarily by central metabolic dysregulation. While the Qalb is a validated index of overall BBB integrity, it does not distinguish among molecule-specific transport mechanisms. Thus, we cannot fully exclude the possibility of differential efflux of QA or KA via active transport. However, these findings reinforce our conclusion that QA-related neurotoxicity in MSA arises from intrinsic CNS metabolic shifts rather than systemic leakage and further support the KYN pathway, particularly KMO activity, as a potential therapeutic target.

Cluster analysis based on the correlation matrix and principal component analysis revealed distinct biomarker groupings in MSA: a metabolically toxic profile defined by QA and 3-HK and a neuroinflammatory and axonal injury marker cluster consisting of sTREM2, GPNMB, and NfL. Different axes between QA and sTREM2 or GPNMB, both established markers of microglial activation, suggesting that QA-related toxicity in MSA may occur independently of or complement classic inflammatory neurotoxic mechanisms. The lack of correlation between QA and inflammatory markers thus further supports the notion that QA elevation in MSA is not merely a consequence of inflammation but rather reflects a shift in central metabolic processes, likely driven by increased KMO activity.

With respect to the relationship between KYN pathway metabolites and clinical symptoms, no significant associations were observed in this study. Previous reports in AD patients indicate that the correlations between cognitive scores and metabolites are generally inconclusive or nonsignificant (Choe *et al*., 2025), whereas studies in PD patients have demonstrated associations between altered KYN metabolism and clinical scores (Jellen *et al*., 2025; Wilson *et al*., 2025). The lack of observed associations with clinical variables in MSA patients, even in the MSA subtype, may reflect their clinical heterogeneity. However, as this was an exploratory study, future hypothesis-driven investigations with adequately powered designs will be necessary to definitively determine whether a true relationship exists.

### Limitations

The cross-sectional design precludes causal inference between biomarker levels and clinical measures. Plasma kynurenine pathway metabolites were not assessed, which limits insights into peripheral-central dynamics; however, given that MSA primarily affects the central nervous system and that QA and KA exhibit poor permeability across the BBB, we focused on CSF as the most appropriate fluid for assessing central kynurenine metabolism. This was a single-center study, and the cohort consisted exclusively of individuals of Japanese ethnicity, which may limit the generalizability of the findings to more diverse populations. Although no significant differences were observed between MSA-C and MSA-P patients, the limited sample size per group restricts conclusive interpretation regarding subtype-specific differences, and larger cohorts are needed to clarify these potential differences. Our study focused on downstream metabolic products and did not measure upstream immunological mediators such as IFN-γ. Thus, IDO1 activation could not be directly assessed; however, the stable KYN/TRP ratio argues against strong IDO1 induction under inflammatory conditions in this cohort. Additionally, the sample size was limited due to the rarity of MSA and the invasive nature of CSF collection. Future longitudinal studies with broader population diversity are warranted to validate these findings.

### Conclusion

This study revealed a pronounced imbalance in KYN pathway metabolites, characterized by elevated levels of QA, reduced levels of KA, and an increased QA-to-KA ratio, which is an indicator of neuroinflammatory processes potentially linked to increased KMO activity. Notably, this shift in TRP-KYN metabolism appeared to be independent of other biomarkers, such as sTREM2, GPNMB, and NfL. Targeting this KMO-associated metabolic imbalance may offer a new metabolic target for MSA.

## Acknowledgments

We thank all the participants for their contributions to this study. This work was supported by the Fujita Mind-Brain Research & Innovation Center for Drug Generation (Fujita Mind-BRIDGe) of Japan’s Peak Research Universities (J-PEAKS) Program funded by the Japan Society for the Promotion of Science (JSPS). This work was also supported by a Grant-in-Aid from the Research Committee of Ataxia, Health Labor Sciences Research Grant, Ministry of Health, Labor and Welfare, Japan.

## Disclosures

The authors declare that there are no conflicts of interest to disclose.

## Data availability statement

The data supporting the findings of this study are available from the corresponding authors upon reasonable request.

## Abbreviations

CSF: cerebrospinal fluid
GPNMB: glycoprotein nonmetastatic melanoma protein B
KA: kynurenic acid
KYN: kynurenine
MSA: multiple system atrophy
NfL: neurofilament light chain
PCA: principal component analysis
QA: quinolinic acid
sTREM2: soluble triggering receptor expressed on myeloid cells 2
SSRI: selective serotonin reuptake inhibitor
TRP: tryptophan
UMSARS: unified MSA rating scale
3-HK: 3-hydroxykynurenine
5-HIAA: 5-hydroxyindoleacetic acid

## Figure legends

**Supplementary Table S1.** Difference between MSA-P and MSA-C.

| Variable | MSA-P | MSA-C | P Value | Adjusted Median Difference<br>(95% CI) |
| --- | --- | --- | --- | --- |
| Tryptophan (μM) | 1.40 [1.17–1.62] | 1.38 [1.22–1.65] | 0.821 | -0.02 (-0.20 to 0.17) |
| 5-HIAA (ng/mL) | 10.9 [5.90–13.2] | 11.4 [7.70–14.7] | 0.366 | -1.41 (-6.62 to 3.41) |
| Kynurenine (nM) | 7.64 [5.55–10.1] | 8.54 [5.74–10.7] | 0.572 | 0.08 (-2.80 to 2.56) |
| 3-Hydroxykynurenine (nM) | 0.11 [0.09–0.34] | 0.17 [0.06–0.26] | 0.695 | 0.05 (-0.02 to 0.19) |
| Kynurenine / Tryptophan ratio | 5.82 [4.25–6.74] | 5.62 [4.24–8.24] | 0.792 | -0.15 (-1.36 to 0.96) |
| Kynurenic acid (nM) | 10.3 [5.51–16.1] | 7.93 [4.69–14.0] | 0.474 | 2.64 (-2.88 to 9.24) |
| Quinolinic acid (nM) | 6.99 [5.09–13.2] | 8.46 [5.50–10.2] | 1.000 | -0.13 (-1.76 to 2.21) |
| QA / KA ratio | 0.76 [0.36–1.24] | 0.85 [0.52–1.69] | 0.486 | -0.07 (-0.82 to 0.49) |
| sTREM2 (pg/mL) | 31830.3 [26535.0–37137.4] | 33051.7 [27117.7–38737.5] | 0.763 | -1082.1 (-5881.7 to 7048.9) |
| GPNMB (pg/mL) | 6937.8 [4614.1–8999.9] | 7027.8 [5250.8–7845.3] | 0.955 | 34.9 (-1473.2 to 1682.9) |
| NfL (pg/mL) | 4934.1 [3466.7–6347.2] | 5342.2 [3473.8–7528.9] | 0.534 | -336.5 (-3097.6 to 1670.8) |

**Supplementary Table S2.** P-values for correlations among biomarkers P-values for correlations among biomarkers, adjusted for age and sex.

|  | Tryptophan | 5-HIAA | Kynurenine | 3-HK | Kynurenic acid | Quinolinic acid | sTREM2 | GPNMB | NfL |
| --- | --- | --- | --- | --- | --- | --- | --- | --- | --- |
| Tryptophan |  | 0.353 | < 0.001 | 0.134 | 0.388 | 0.220 | 0.502 | 0.099 | 0.119 |
| 5-HIAA | 0.353 |  | 0.721 | 0.815 | 0.150 | 0.684 | 0.523 | 0.916 | 0.923 |
| Kynurenine | < 0.001 | 0.721 |  | 0.002 | 0.709 | < 0.001 | 0.336 | 0.697 | 0.898 |
| 3-HK | 0.134 | 0.815 | 0.002 |  | 0.363 | 0.020 | 0.833 | 0.712 | 0.672 |
| Kynurenic acid | 0.388 | 0.150 | 0.709 | 0.363 |  | 0.886 | 0.256 | 0.051 | 0.952 |
| Quinolinic acid | 0.220 | 0.684 | < 0.001 | 0.020 | 0.886 |  | 0.678 | 0.871 | 0.997 |
| sTREM2 | 0.502 | 0.523 | 0.336 | 0.833 | 0.256 | 0.678 |  | < 0.001 | 0.011 |
| GPNMB | 0.099 | 0.916 | 0.697 | 0.712 | 0.051 | 0.871 | < 0.001 |  | < 0.001 |
| NfL | 0.119 | 0.923 | 0.898 | 0.672 | 0.952 | 0.997 | 0.011 | < 0.001 |  |

|  | Tryptophan | 5-HIAA | Kynurenine | 3-HK | Kynurenic acid | Quinolinic acid | sTREM2 | GPNMB | NfL |
| --- | --- | --- | --- | --- | --- | --- | --- | --- | --- |
| Tryptophan |  | 0.798 | < 0.001 | 0.048 | 0.582 | 0.213 | 0.580 | 0.589 | 0.509 |
| 5-HIAA | 0.798 |  | 0.986 | 0.788 | 0.093 | 0.974 | 0.345 | 0.771 | 0.611 |
| Kynurenine | < 0.001 | 0.986 |  | < 0.001 | 0.740 | < 0.001 | 0.242 | 0.997 | 0.501 |
| 3-HK | 0.048 | 0.788 | < 0.001 |  | 0.516 | 0.002 | 0.993 | 0.795 | 0.875 |
| Kynurenic acid | 0.582 | 0.093 | 0.740 | 0.516 |  | 0.705 | 0.434 | 0.079 | 0.862 |
| Quinolinic acid | 0.213 | 0.974 | < 0.001 | 0.002 | 0.705 |  | 0.390 | 0.601 | 0.524 |
| sTREM2 | 0.580 | 0.345 | 0.242 | 0.993 | 0.434 | 0.390 |  | < 0.001 | 0.004 |
| GPNMB | 0.589 | 0.771 | 0.997 | 0.795 | 0.079 | 0.601 | < 0.001 |  | 0.003 |
| NfL | 0.509 | 0.611 | 0.501 | 0.875 | 0.862 | 0.524 | 0.004 | 0.003 |  |

These p-values correspond to the Spearman $\rho$ values presented in Supplementary Figure S1A.

**Supplementary Table S3.** P-values for correlations between biomarkers and clinical scores P-values for correlations among biomarkers, adjusted for age and sex.

|  | Tryptophan | 5-HIAA | Kynurenine | 3-HK | Kynurenic acid | Quinolinic acid | sTREM2 | GPNMB | NfL |
| --- | --- | --- | --- | --- | --- | --- | --- | --- | --- |
| Duration | 0.033 | 0.010 | 0.536 | 0.339 | 0.101 | 0.918 | 0.254 | 0.452 | 0.180 |
| UMSARS I | 0.201 | 0.003 | 0.216 | 0.999 | 0.559 | 0.713 | 0.887 | 0.693 | 0.864 |
| UMSARS II | 0.110 | 0.005 | 0.217 | 0.418 | 0.331 | 0.851 | 0.635 | 0.875 | 0.917 |
| UMSARS IV | 0.075 | 0.006 | 0.098 | 0.782 | 0.939 | 0.937 | 0.530 | 0.818 | 0.593 |
| MMSE | 0.459 | 0.242 | 0.632 | 0.625 | 0.100 | 0.532 | 0.589 | 0.716 | 0.077 |
| MoCA-J | 0.860 | 0.397 | 0.485 | 0.885 | 0.865 | 0.817 | 0.241 | 0.794 | 0.177 |
| ACE-R | 0.646 | 0.030 | 0.219 | 0.320 | 0.129 | 0.469 | 0.664 | 0.637 | 0.104 |
| FAB | 0.452 | 0.260 | 0.810 | 0.543 | 0.436 | 0.835 | 0.973 | 0.654 | 0.367 |
| GDS | 0.975 | 0.686 | 0.310 | 0.415 | 0.451 | 0.287 | 0.033 | 0.010 | 0.015 |
| SCOPA-AUT | 0.515 | < 0.001 | 0.549 | 0.219 | 0.775 | 0.708 | 0.825 | 0.637 | 0.662 |

|  | Tryptophan | 5-HIAA | Kynurenine | 3-HK | Kynurenic acid | Quinolinic acid | sTREM2 | GPNMB | NfL |
| --- | --- | --- | --- | --- | --- | --- | --- | --- | --- |
| Duration | 0.069 | 0.008 | 0.300 | 0.503 | 0.062 | 0.473 | 0.307 | 0.265 | 0.593 |
| UMSARS I | 0.528 | 0.003 | 0.476 | 0.771 | 0.422 | 0.591 | 0.734 | 0.994 | 0.593 |
| UMSARS II | 0.352 | 0.007 | 0.613 | 0.351 | 0.250 | 0.638 | 0.760 | 0.744 | 0.729 |
| UMSARS IV | 0.163 | 0.009 | 0.262 | 0.723 | 0.930 | 0.823 | 0.577 | 0.690 | 0.893 |
| MMSE | 0.633 | 0.295 | 0.719 | 0.953 | 0.108 | 0.595 | 0.737 | 0.825 | 0.149 |
| MoCA-J | 0.806 | 0.473 | 0.129 | 0.502 | 0.761 | 0.097 | 0.167 | 0.784 | 0.316 |
| ACE-R | 0.999 | 0.046 | 0.026 | 0.167 | 0.099 | 0.037 | 0.490 | 0.780 | 0.229 |
| FAB | 0.591 | 0.407 | 0.480 | 0.618 | 0.442 | 0.097 | 0.988 | 0.470 | 0.572 |
| GDS | 0.561 | 0.607 | 0.121 | 0.457 | 0.349 | 0.121 | 0.031 | 0.033 | 0.004 |
| SCOPA-AUT | 0.913 | < 0.001 | 0.824 | 0.402 | 0.653 | 0.614 | 0.701 | 0.511 | 0.726 |

These p-values correspond to the Spearman $\rho$ values presented in Supplementary Figure S1B.

**Supplementary Figure S1.**
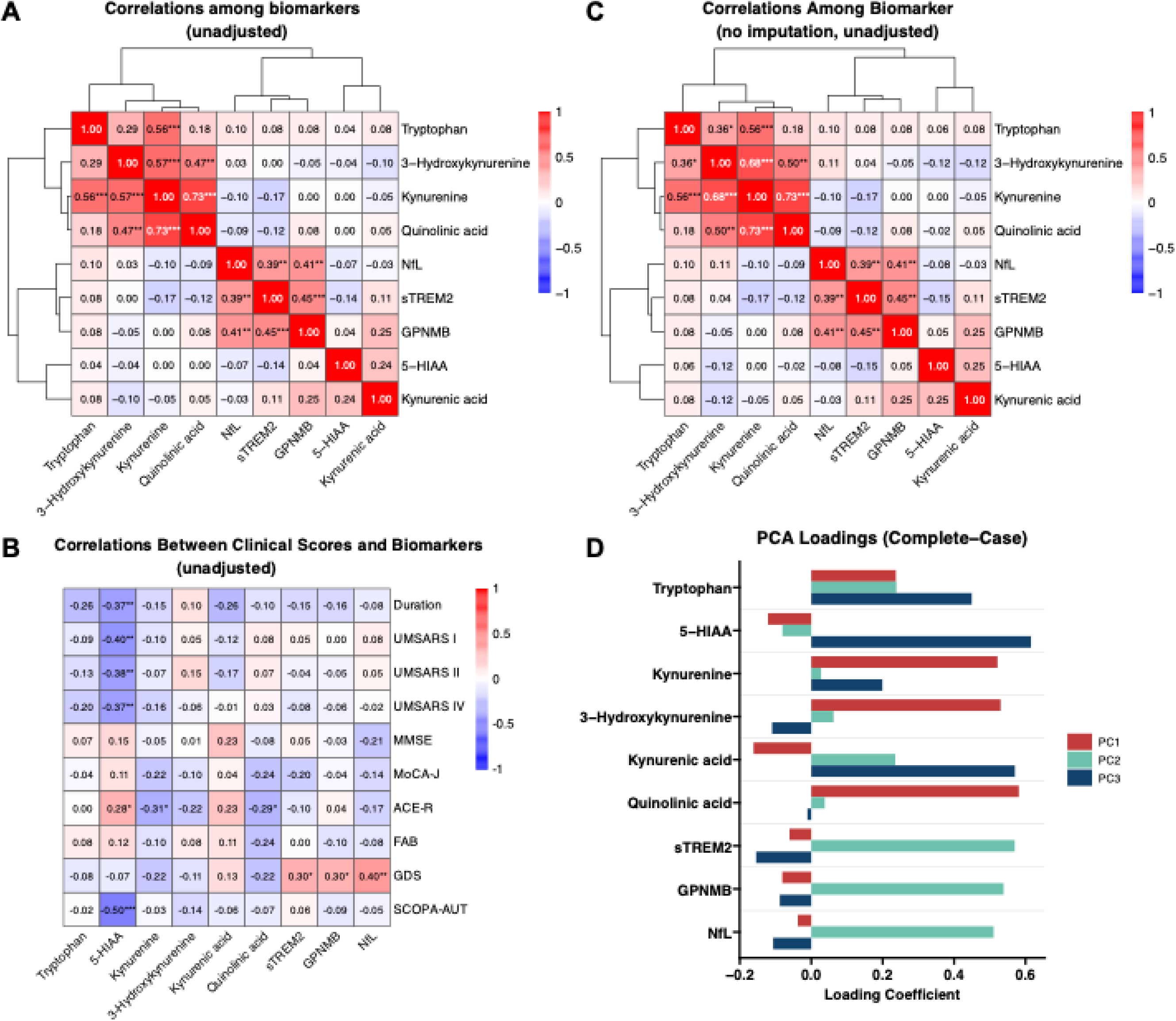
Sensitivity analysis of relationship among biomarkers and clinical scores. A. Correlations among biomarkers unadjusted for age and sex. Values represent Spearman’s ρ. * p < 0.05; ** p < 0.01; *** p < 0.001. B. Correlations between clinical scores and biomarkers, unadjusted for age and sex. C. Correlations among biomarkers adjusted for age and sex using pairwise deletion without multiple imputation. Values represent Spearman’s ρ. * p < 0.05; ** p < 0.01; *** p < 0.001. D. Principal component analysis (PCA) of biomarkers showing loadings for principal components (PCs) 1 to 3, using listwise deletion without imputation. 5-HIAA, 5-hydroxyindoleacetic acid; GPNMB, glycoprotein nonmetastatic melanoma protein B; NfL, neurofilament light chain; sTREM2, soluble triggering receptor expressed on myeloid cells 2.

**Supplementary Figure S2.**
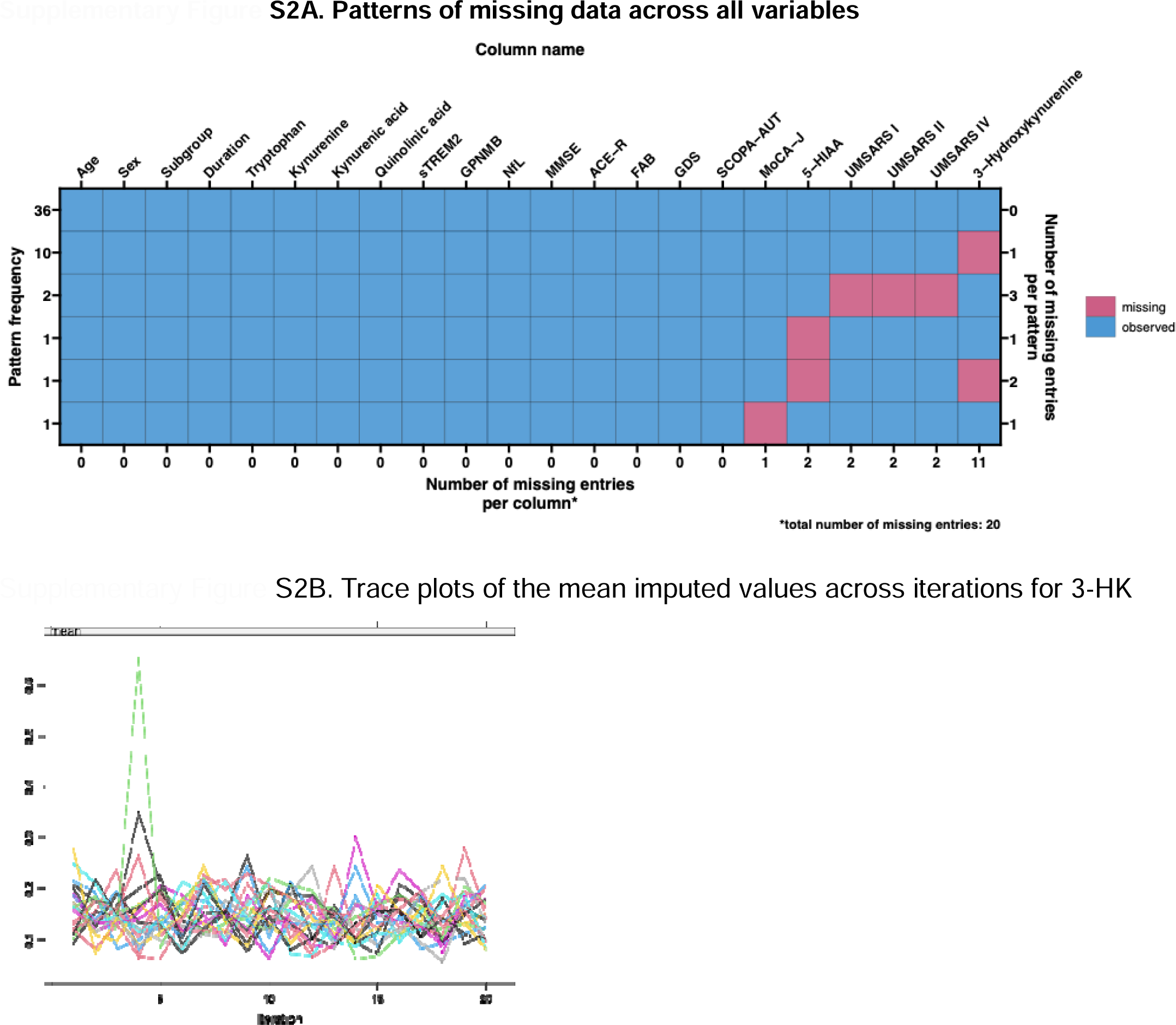
A illustrates the patterns of missing data across all variables, highlighting both variable-wise and case-wise missingness. Among 51 patients with MSA, missing values were observed in 3-HK (11 cases, 22%), 5-HIAA (2 cases, 3.9%), UMSARS part I and II (2 cases, 3.9%), part IV (2 cases, 3.9%), and MoCA-J (1 case, 2.0%). Given the relatively high missingness in 3-HK, this variable was evaluated the convergence of the multiple imputation model ad iteration 20. Trace plots of the mean imputed values across iterations for 3-HK are presented in Supplementary Figure S2B (shown for a subset of 20 imputations due to plot density). In addition, the potential scale reduction factor (PSRF) for 3-HK was calculated to confirm convergence, with values falling below 1.1, indicating acceptable stability of the imputations (Huang and Keller 2025). Huang, Francis, and Brian Keller. 2025. “Working with Missing Data in Large-Scale Assessments.” *Large-Scale Assessments in Education* 13 (1). https://doi.org/10.1186/s40536-025-00248-9.

**Supplementary Figure S3.**
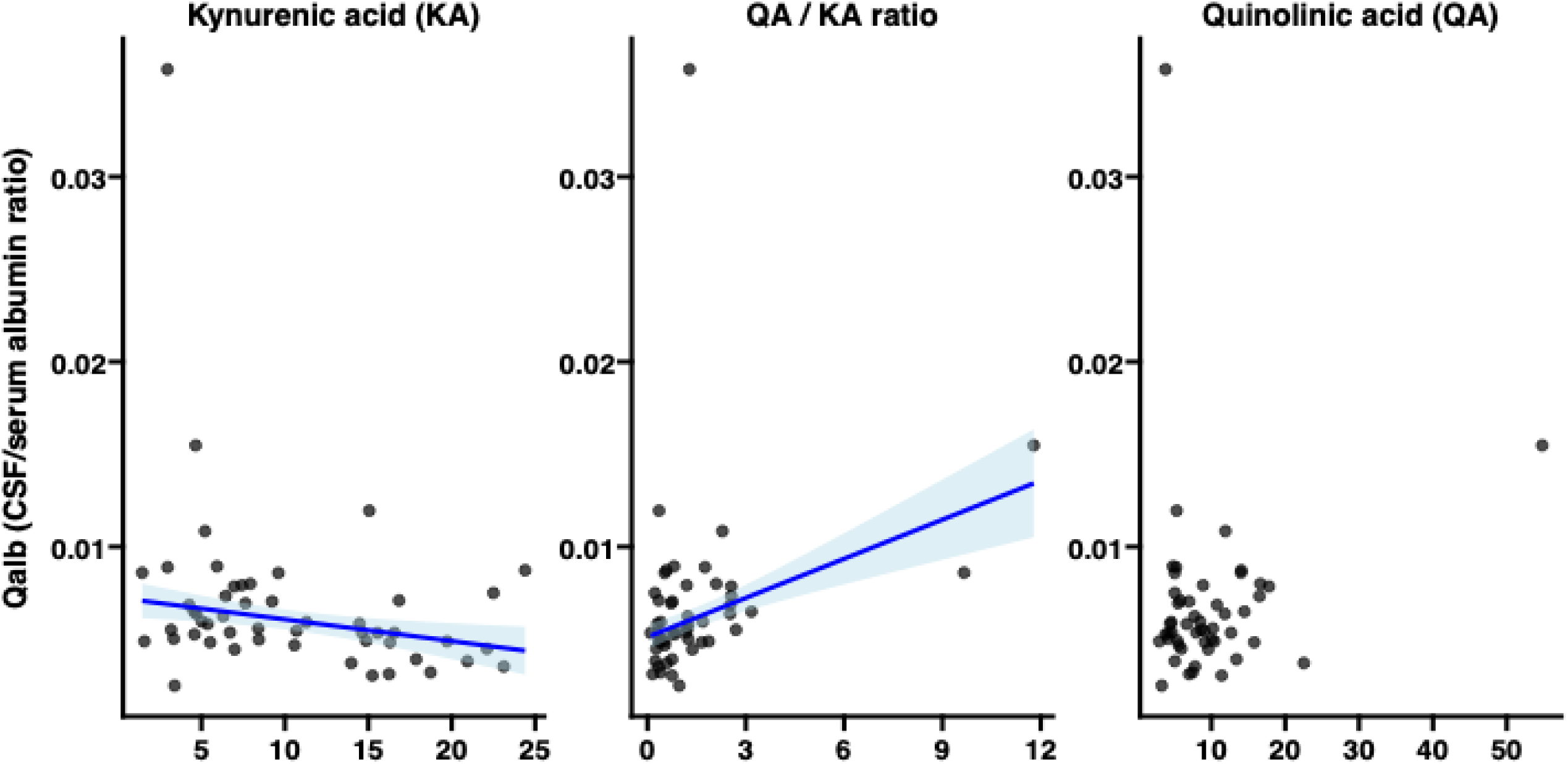
Correlation between Qalb and KA, QA, and the QA/KA ratio. To evaluate the relationships between Qalb and key kynurenine metabolites, we computed Spearman’s rank correlation coefficients between Qalb and kynurenic acid (KA), quinolinic acid (QA), and the QA/KA ratio. Robust linear regression was applied to visualize the trend lines with 95% confidence intervals, limited to cases where the correlation reached statistical significance (p < 0.05). Further details are provided in the main text.

## Notes

### Competing Interest Statement

The authors have declared no competing interest.

### Author Declarations

The Ethics Committee of Fujita Health University Hospital approved this study (approval number: HM23-296).

## References

Badawy AA-B, Guillemin GJ. Species differences in tryptophan metabolism and disposition. Int J Tryptophan Res 2022; 15: 11786469221122512.

Bender DA. Biochemistry of tryptophan in health and disease. Mol Aspects Med 1983; 6: 101–97.

Castellano-Gonzalez G, Jacobs KR, Don E, Cole NJ, Adams S, Lim CK, et al. Kynurenine 3-monooxygenase activity in human primary neurons and effect on cellular bioenergetics identifies new neurotoxic mechanisms. Neurotox Res 2019; 35: 530–41.

Choe K, Bakker L, van den Hove DLA, Eussen SJPM, Kenis G, Ramakers IHGB, et al. Kynurenine pathway dysregulation in cognitive impairment and dementia: a systematic review and meta-analysis [Internet]. GeroScience 2025 Available from: 10.1007/s11357-025-01636-3

Gabdulkhaev R, Shimizu H, Kanazawa M, Kuroha Y, Hasegawa A, Idezuka J, et al. Blood-brain barrier dysfunction in multiple system atrophy: A human postmortem study. Neuropathology 2025; 45: 210–22.

van Ginkel JR. Handling missing data in principal component analysis using multiple imputation. In: Methodology of Educational Measurement and Assessment. Cham: Springer International Publishing; 2023. p. 141–61

Haq S, Grondin JA, Khan WI. Tryptophan-derived serotonin-kynurenine balance in immune activation and intestinal inflammation. FASEB J 2021; 35: e21888.

Hasegawa M, Kunisawa K, Wulaer B, Kubota H, Kurahashi H, Sakata T, et al. Chronic stress induces behavioural changes through increased kynurenic acid by downregulation of kynurenine-3-monooxygenase with microglial decline. Br J Pharmacol 2025; 182: 1466–86.

Hoffmann A, Ettle B, Battis K, Reiprich S, Schlachetzki JCM, Masliah E, et al. Oligodendroglial α-synucleinopathy-driven neuroinflammation in multiple system atrophy. Brain Pathol 2019; 29: 380–96.

Huang Y, Zhao M, Chen X, Zhang R, Le A, Hong M, et al. Tryptophan metabolism in central nervous system diseases: Pathophysiology and potential therapeutic strategies. Aging Dis 2023; 14: 858–78.

Hughes TD, Güner OF, Iradukunda EC, Phillips RS, Bowen JP. The kynurenine pathway and kynurenine 3-monooxygenase inhibitors. Molecules 2022; 27: 273.

Javors MA, Bowden CL, Maas JW. 3-methoxy-4-hydroxyphenylglycol, 5-hydroxyindoleacetic acid, and homovanillic acid in human cerebrospinal fluid. Storage and measurement by reversed-phase high-performance liquid chromatography and coulometric detection using 3-methoxy-4-hydroxyphenyllactic acid as an internal standard. J Chromatogr 1984; 336: 259–69.

Jellen LC, Escobar Galvis ML, Sha Q, Isaguirre C, Johnson A, Madaj Z, et al. Sex differences in peripheral and central dysregulation of the kynurenine pathway in Parkinson’s disease. NPJ Parkinsons Dis 2025; 11: 116.

Kaleta M, Hényková E, Menšíková K, Friedecký D, Kvasnička A, Klíčová K, et al. Patients with neurodegenerative proteinopathies exhibit altered tryptophan metabolism in the serum and cerebrospinal fluid. ACS Chem Neurosci 2024; 15: 582–92.

Kawabata K, Krismer F, Ito M, Hara K, Bagarinao E, Beliveau V, et al. Trajectories of pontine volume in patients with multiple system atrophy [Internet]. Mov Disord 2025[cited 2025 Apr 23] Available from: 10.1002/mds.30182

Krismer F, Péran P, Beliveau V, Seppi K, Arribarat G, Pavy-Le Traon A, et al. Progressive Brain Atrophy in Multiple System Atrophy: A Longitudinal, Multicenter, Magnetic Resonance Imaging Study [Internet]. Mov Disord 2023 Available from: 10.1002/mds.29633

Leńska-Mieciek M, Madetko-Alster N, Alster P, Królicki L, Fiszer U, Koziorowski D. Inflammation in multiple system atrophy. Front Immunol 2023; 14: 1214677.

Lugo-Huitrón R, Ugalde Muñiz P, Pineda B, Pedraza-Chaverrí J, Ríos C, Pérez-de la Cruz V. Quinolinic acid: an endogenous neurotoxin with multiple targets. Oxid Med Cell Longev 2013; 2013: 104024.

Mailman RB, Kilts CD. Analytical considerations for quantitative determination of serotonin and its metabolically related products in biological matrices. Clin Chem 1985; 31: 1849–54.

Marshall A, Altman DG, Holder RL, Royston P. Combining estimates of interest in prognostic modelling studies after multiple imputation: current practice and guidelines. BMC Med Res Methodol 2009; 9: 57.

Mor A, Tankiewicz-Kwedlo A, Krupa A, Pawlak D. Role of kynurenine pathway in oxidative stress during neurodegenerative disorders. Cells 2021; 10: 1603.

Mori Y, Mouri A, Kunisawa K, Hirakawa M, Kubota H, Kosuge A, et al. Kynurenine 3-monooxygenase deficiency induces depression-like behavior via enhanced antagonism of α7 nicotinic acetylcholine receptors by kynurenic acid. Behav Brain Res 2021; 405: 113191.

Moroni F, Cozzi A, Sili M, Mannaioni G. Kynurenic acid: a metabolite with multiple actions and multiple targets in brain and periphery. J Neural Transm (Vienna) 2012; 119: 133–9.

Nagao R, Mizutani Y, Shima S, Ueda A, Ito M, Yoshimoto J, et al. Correlations between serotonin impairments and clinical indices in multiple system atrophy. Eur J Neurol 2024; 31: e16158.

Okuda S, Nishiyama N, Saito H, Katsuki H. 3-Hydroxykynurenine, an endogenous oxidative stress generator, causes neuronal cell death with apoptotic features and region selectivity. J Neurochem 1998; 70: 299–307.

Parrott JM, O’Connor JC. Kynurenine 3-monooxygenase: An influential mediator of neuropathology. Front Psychiatry 2015; 6: 116.

Pedersen AB, Mikkelsen EM, Cronin-Fenton D, Kristensen NR, Pham TM, Pedersen L, et al. Missing data and multiple imputation in clinical epidemiological research. Clin Epidemiol 2017; 9: 157–66.

Platten M, Nollen EAA, Röhrig UF, Fallarino F, Opitz CA. Tryptophan metabolism as a common therapeutic target in cancer, neurodegeneration and beyond. Nat Rev Drug Discov 2019; 18: 379–401.

Rubin DB. Multiple Imputation for Nonresponse in Surveys. 99th ed. Nashville, TN: John Wiley & Sons; 1987

Schwarcz R, Bruno JP, Muchowski PJ, Wu H-Q. Kynurenines in the mammalian brain: when physiology meets pathology. Nat Rev Neurosci 2012; 13: 465–77.

Sorgdrager FJH, Vermeiren Y, Van Faassen M, van der Ley C, Nollen EAA, Kema IP, et al. Age- and disease-specific changes of the kynurenine pathway in Parkinson’s and Alzheimer’s disease. J Neurochem 2019; 151: 656–68.

van der Velpen V, Teav T, Gallart-Ayala H, Mehl F, Konz I, Clark C, et al. Systemic and central nervous system metabolic alterations in Alzheimer’s disease. Alzheimers Res Ther 2019; 11: 93.

Vieira BDM, Radford RA, Chung RS, Guillemin GJ, Pountney DL. Neuroinflammation in multiple system atrophy: Response to and cause of α-synuclein aggregation. Front Cell Neurosci 2015; 9: 437.

Wakabayashi K, Yoshimoto M, Tsuji S, Takahashi H. Alpha-synuclein immunoreactivity in glial cytoplasmic inclusions in multiple system atrophy. Neurosci Lett 1998; 249: 180–2.

Wang Y, Zhang Y, Wang W, Zhang Y, Dong X, Liu Y. Diverse physiological roles of kynurenine pathway metabolites: Updated implications for health and disease. Metabolites 2025; 15: 210.

Watanabe H, Saito Y, Terao S, Ando T, Kachi T, Mukai E, et al. Progression and prognosis in multiple system atrophy: an analysis of 230 patients. Brain 2002; 125: 1070–83.

Wenning GK, Stankovic I, Vignatelli L, Fanciulli A, Calandra-Buonaura G, Seppi K, et al. The Movement Disorder Society Criteria for the Diagnosis of Multiple System Atrophy. Mov Disord 2022; 37: 1131–48.

Wilson EN, Umans J, Swarovski MS, Minhas PS, Mendiola JH, Midttun Ø, et al. Parkinson’s disease is characterized by vitamin B6-dependent inflammatory kynurenine pathway dysfunction. NPJ Parkinsons Dis 2025; 11: 96.

